# SEROPREVALENCE AND CLINICAL SPECTRUM OF SARS-CoV-2 INFECTION IN THE FIRST VERSUS THIRD TRIMESTER OF PREGNANCY

**DOI:** 10.1101/2020.06.17.20134098

**Authors:** Francesca Crovetto, Fàtima Crispi, Elisa Llurba, Francesc Figueras, María Dolores Gómez-Roig, Eduard Gratacós

## Abstract

**Introduction:** Case registries of pregnant women diagnosed with coronavirus disease (COVID-19) by polymerase chain reaction (PCR) have reported that the majority experienced mild infection, but up to 9% may require critical care.^1^ Most COVID-19 cases published were in the third trimester of pregnancy, which could reflect reporting bias, higher risk of infection or increased disease severity in late pregnancy.^2^ Seroprevalence studies may allow reliable estimates of the susceptibility to infection and clinical spectrum since they include asymptomatic and mild infections not tested for PCR. We evaluated the seroprevalence and clinical presentation of severe acute respiratory syndrome coronavirus 2 (SARS-CoV-2) infection in pregnant women in the first and third trimester.

**Methods:** The study was approved by the Institutional Review Board at each institution and informed consent was obtained. We recruited 874 consecutive pregnancies attending for first trimester screening (10-16 weeks’ gestation, n=372) or delivery (n=502) from April 14 to May 5. All women were interviewed with a structured questionnaire for COVID-19 symptoms two months prior to sampling. SARS-CoV-2 IgG and IgM/IgA antibodies were tested (COVID-19 VIRCLIA® Monotest, Vircell Microbiologist, Spain; reported sensitivity 70% IgG and 89% IgM/IgA, and specificity 89% and 99% respectively). Indeterminate results were re-tested (VITROS® Immunodiagnostic Products Anti-SARS-CoV2 Total Tests, Ortho Clinical Diagnostics, USA; 100% sensitivity and specificity) and re-classified as positive or negative. Women with COVID-19 were diagnosed and managed according to standard protocols and guidelines^3,4^. Statistical differences were tested using the *χ*^2^ test or Student t-test as appropriate (p<0.05).

**Results:** A total of 125 of 874 women (14.3%) were positive for either IgG or IgM/IgA SARS-CoV-2 antibodies, 54/372 (14.5%) in the first and 71/502 (14.1%) in the third trimester. A total of 75/125 (60%) reported no symptoms of COVID-19 in the past 2 months, whereas 44 (35.2%) reported one or more symptoms, of which 31 (24.8%) had at least 3 symptoms or anosmia and 8 (6.4%) dyspnea. Overall, 7 women (5.6%) were admitted for persistent fever (>38°) despite paracetamol and dyspnea, of which 3 had signs of pneumonia on chest radiography. All 3 had criteria for severity (bilateral chest condensation, respiratory rate>30 and leukopenia) and required oxygen support but not critical care or mechanical ventilation, and they were all discharged well. The rates of symptomatic infection, hospital admission or dyspnea were significantly higher in third trimester women (Table and Figure).

**Discussion:** The 14.3% seroprevalence of SARS-COV-2 in pregnant women in this study was substantially larger than the contemporary rates of PCR positive cases (0.78%) reported for women 20-40y in Barcelona.^5^ The data confirm that COVID-19 is asymptomatic in the majority of pregnant women^6^ and illustrate the value of seroprevalence studies to capture the high proportion of asymptomatic or mild infections. In this study, none of the 125 pregnant women with SARS-CoV-2 infection required critical care as compared to 9% reported in cases diagnosed with PCR.^1^ However, the proportion of infections with symptoms or dyspnea was remarkably higher in the third trimester, and these results are in line with COVID-19 registries, reporting that 81% of hospitalized women were in late pregnancy or peripartum.^1^

These results provide reassuring information that, even in settings with a high prevalence, SARS-CoV-2 infection in pregnancy mostly presents with asymptomatic or mild clinical forms. The susceptibility to infection seemed to be the same in the first and the third trimesters of gestation. The data further suggest that, as with other respiratory viruses, COVID-19 could be more severe and require increased surveillance in late pregnancy. These findings should be confirmed and extended with larger consecutive prevalence studies in pregnancy.

## Introduction

Case registries of pregnant women diagnosed with coronavirus disease (COVID-19) by polymerase chain reaction (PCR) have reported that the majority experienced mild infection, but up to 9% may require critical care.^1^ Most COVID-19 cases published were in the third trimester of pregnancy, which could reflect reporting bias, higher risk of infection or increased disease severity in late pregnancy.^2^ Seroprevalence studies may allow reliable estimates of the susceptibility to infection and clinical spectrum since they include asymptomatic and mild infections not tested for PCR. We evaluated the seroprevalence and clinical presentation of severe acute respiratory syndrome coronavirus 2 (SARS-CoV-2) infection in pregnant women in the first and third trimester.

## Methods

The study was approved by the Institutional Review Board at each institution and informed consent was obtained. We recruited 874 consecutive pregnancies attending for first trimester screening (10-16 weeks’ gestation, n=372) or delivery (n=502) from April 14 to May 5. All women were interviewed with a structured questionnaire for COVID-19 symptoms two months prior to sampling. SARS-CoV-2 IgG and IgM/IgA antibodies were tested (COVID-19 VIRCLIA® Monotest, Vircell Microbiologist, Spain; reported sensitivity 70% IgG and 89% IgM/IgA, and specificity 89% and 99% respectively). Indeterminate results were re-tested (VITROS® Immunodiagnostic Products Anti-SARS-CoV2 Total Tests, Ortho Clinical Diagnostics, USA; 100% sensitivity and specificity) and re-classified as positive or negative. Women with COVID-19 were diagnosed and managed according to standard protocols and guidelines^3,4^. Statistical differences were tested using the *χ*^2^ test or Student t-test as appropriate (p<0.05).

## Results

A total of 125 of 874 women (14.3%) were positive for either IgG or IgM/IgA SARS-CoV-2 antibodies, 54/372 (14.5%) in the first and 71/502 (14.1%) in the third trimester. A total of 75/125 (60%) reported no symptoms of COVID-19 in the past 2 months, whereas 44 (35.2%) reported one or more symptoms, of which 31 (24.8%) had at least 3 symptoms or anosmia and 8 (6.4%) dyspnea. Overall, 7 women (5.6%) were admitted for persistent fever (>38°) despite paracetamol and dyspnea, of which 3 had signs of pneumonia on chest radiography. All 3 had criteria for severity (bilateral chest condensation, respiratory rate>30 and leukopenia) and required oxygen support but not critical care or mechanical ventilation, and they were all discharged well. The rates of symptomatic infection, hospital admission or dyspnea were significantly higher in third trimester women (Table and Figure).

**Table.** Baseline characteristics and clinical features of pregnant women positive for SARS-CoV-2 infection (N=125) in the first versus third trimester.

| Characteristics | First Trimester<br>N=54 | Third Trimester<br>N=71 | p |
| --- | --- | --- | --- |
| Age (years) | 33.7 (5.1) | 31.8 (6.1) | 0.062 |
| White ethnicity | 31 (57.4) | 43 (60.6) | 0.697 |
| Body mass index (Kg/m<sup>2</sup>) | 24.1 (6.2) | 23.9 (4.3) | 0.209 |
| Smoking status | 4 (7.4) | 6 (8.5) | 0.543 |
| Medical history |  |  |  |
| Chronic hypertension | 1 (1.9) | 2 (2.8) | 0.602 |
| Diabetes mellitus | 1 (1.9) | 0 (0) | 0.432 |
| Autoimmune disease | 2 (3.7) | 0 (0) | 0.185 |
| Respiratory disease | 2 (3.7) | 3 (4.2) | 0.297 |
| Nulliparous | 34 (63) | 36 (50.7) | 0.095 |
| Multiple pregnancy | 2 (3.7) | 1 (1.4) | 0.398 |
| Symptoms compatible with COVID-19 within last 2 months |  |  |  |
| None | 38 (70.4) | 37 (52.1) | 0.012 |
| Fever | 7 (13) | 16 (22.5) | 0.152 |
| Dry cough | 5 (9.3) | 17 (23.9) | 0.034 |
| Dyspnea | 0 (0) | 8 (11.3) | 0.010 |
| Tiredness | 5 (9.3) | 4 (5.6) | 0.307 |
| Sore throat | 6 (11.1) | 3 (4.2) | 0.116 |
| Rash on skin, or discoloration of fingers/toes | 0 (0) | 0 (0) | NA |
| Diarrhea | 4 (7.4) | 5 (7) | 0.575 |
| Loss of taste or smell | 7 (13) | 16 (22.5) | 0.152 |
| At least 3 symptoms or anosmia | 9 (16.7) | 22 (31.0) | 0.065 |
| Fever, cough and dyspnea | 0 (0) | 8 (11.3) | 0.010 |
| Hospital admission for COVID-19 symptoms | 0 (0) | 7 (9.9) | 0.019 |

SARS-CoV-2: severe acute respiratory syndrome coronavirus 2
Data are expressed as n (%) or mean (SD)

**Figure.**
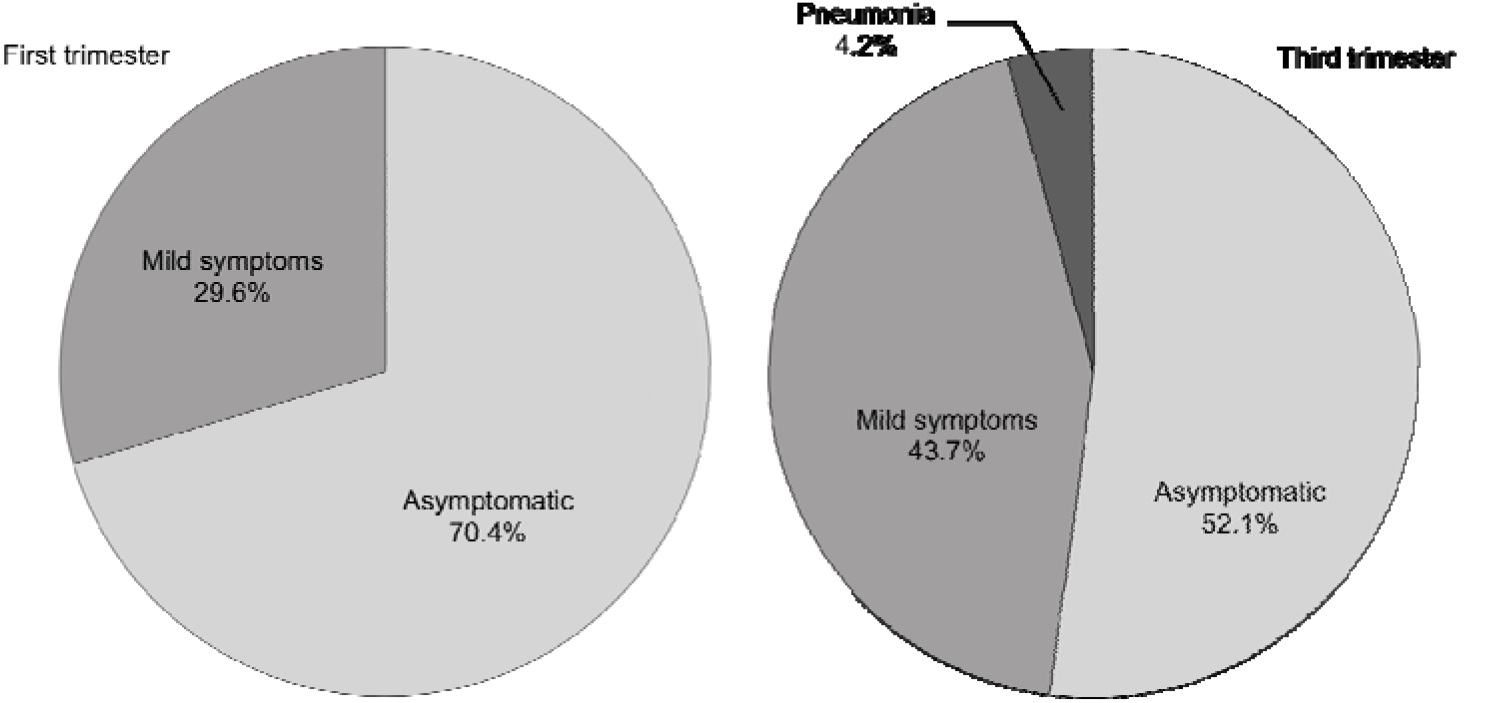
Clinical spectrum of SARS-CoV-2 infection in pregnant women in the first and third trimester.

## Discussion

The 14.3% seroprevalence of SARS-COV-2 in pregnant women in this study was substantially larger than the contemporary rates of PCR positive cases (0.78%) reported for women 20-40y in Barcelona.^5^ The data confirm that COVID-19 is asymptomatic in the majority of pregnant women^6^ and illustrate the value of seroprevalence studies to capture the high proportion of asymptomatic or mild infections. In this study, none of the 125 pregnant women with SARS-CoV-2 infection required critical care as compared to 9% reported in cases diagnosed with PCR.^1^ However, the proportion of infections with symptoms or dyspnea was remarkably higher in the third trimester, and these results are in line with COVID-19 registries, reporting that 81% of hospitalized women were in late pregnancy or peripartum.^1^

## Data Availability

We state that all data will be available if needed.

## Additional contributions

We thank all the medical staff, residents, midwifes and nurses of BCNatal (Hospitals Sant Joan de Déu and Clínic) and Hospital Sant Pau, especially Rosalia Pascal, MD, Marta Larroya, MD, Cristina Trilla, MD, Martí Cantallops, MD, Marta Camacho, MD, M. Carmen Medina, MD, Irene Casas, MD, Marta Tortajada, MD, Àlex Cahuana, MD, Patricia Muro, MD, Marta Valdés, MD, David Boada, MD and Anna Mundo, MD, for careful data collection; Angela Arranz, PhD, for her key role in the coordination of the nursing research team; Imma Mercade, PhD, Elena Casals, PhD, and Josefina Mora, PhD, for their contribution in the collection of first trimester samples; Jordi Yague, PhD, and M. Angeles Marcos, PhD, for supervising the antibodies analyses; and the Biobanks of Fundació Sant Joan de Déu, Clínic-IDIBAPS and Sant Pau for valuable management of samples. None of them received any compensation for their contribution.

